# Challenging the Chronic Perverse Bias in Prosthetic Valve Design: A Pathway Opens for Advanced Mechanical Valves

**DOI:** 10.1101/2024.09.12.24313331

**Authors:** Lawrence N. Scotten, Dylan Goode, Rolland Siegel, David J. Blundon, James W. Dutton, Hadi Mohammadi

## Abstract

**Objective:**

- In vitro evaluation of several prototype mechanical valves compared to present-day controls.

**Method:**

- simulated normal cardiac pressures and flows
- gravity pressure head column tester flows.
- recorded valve hydrodynamics and kinematics

**Results:**

- valves superior in performance to clinical controls

**Conclusions:**

- Prototype MHV candidates outperform the closing performance of present day SAVR prosthetic valves, including bioprosthetic control.

**Competing Interests:** None declared

**Financial Disclosure:** This research was performed on a pro bono basis by indicted coauthors*, in part, to assist coauthor and Ph.D. candidate Dylan Goode.

**CENTRAL MESSAGE:** In-vitro dynamics of optimized prototype mechanical heart valves (MHVs) outperformed those of current clinical prosthetic SAVR valves. -Achieving the objective of an anticoagulation-free and durable MHV is directly related to mitigation of detrimental valve closing hydrodynamics and kinematics.

**PERSPECTIVE:** Improvements in prosthetic valve performance and durability notwithstanding, the objective of an anti-coagulation free device with durability exceeding the projected life expectancy of all recipients has not been achieved. Our work identifies a design and development void that may have significantly delayed progress toward this objective.

**SIGNIFICANCE:** Our results challenge a longstanding bias in valve design, shifting the focus towards crucial behavior during valve closure. This study paves the way for advanced mechanical valves bringing us closer to the elusive goal of anticoagulation-free performance—a long-awaited milestone in the evolution of prosthetic valves.

## INTRODUCTION AND BACKGROUND

From the earliest commercially prepared ball valves by Starr and Harken, single disk occluder models from Shiley (1971), (Kaster et al. 1969) and Cromie (Magovern GJ, Cromie HW. 1963) adaptation of pyrolytic carbon from nuclear power application to heart valve components by Bokros (Bokros JC. 1989) led to innovative bi-leaflet valves by Nicoloff and Hansen. Throughout this long developmental phase, valve designers focused on extending durability, improving hemodynamic performance related to clinical gradients and reducing thrombogenicity through the modification of prosthetic surface-blood contact interactions. The iconic example of this design focus is the St. Jude Medical (SJM) valve. This prosthesis has two pivoting occluders retained by leaflet-tabs within butterfly shaped recesses in the housing. Nicoloff first implanted the SJM valve on October 3, 1977 (Nicoloff DM 1981) and currently has >3 million implants as reported by Copilot AI. However, while valve designers focused on minimizing forward flow energy losses to reduce clinical pressure gradient, our prior work exposed a persistent bias during 60+ years of valve development which ignored crucial events near or at valve closure that contribute to or are responsible for shear damage to formed blood components. In this study, we utilize our Leonardo modified pulse duplicator (Scotten et al., 2024) to compare closure dynamics in contemporary bi-leaflet and pericardial valves with long clinical histories to an innovative tri-leaflet mechanical valve from (Perrier 2023), (Carrel et al. 2023), an experimental design by (Mohammadi, 2023) and (Goode 2024) intended to achieve long-term anticoagulation independence and an experimental bi-leaflet valve BV3D (Scotten and Siegel, 2015), crafted in our laboratory.

## METHODS

### Pulse Duplicator Experiments

#### Assessment of dynamic flow velocity

The Leonardo system enables the assessment of dynamic flow velocity using an electromagnetic magnetic flow probe and photodiode measurement of Projected Open Area (POA). We equated flow velocity as:

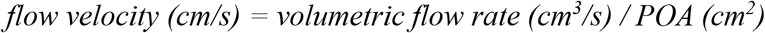

Assumptions and considerations about the POA region are that the flow velocity profile is uniform, however, this may not be valid when valve occluder motion and volumetric flowrate are irregular. Consistent with Bernoulli’s principle, our experiments confirmed that as POA decreases, valve flow velocity increases and, conversely, decreases when POA increases.

The dynamic flow velocity in cm/s is the ratio of two measured quantities, -volumetric flow rate from a calibrated electromagnetic flow probe in cm^3^s^-1^/ Projected Valve Area (POA) in cm^2^. We measured signals in two locations, the electromagnetic flow probe acquired the pulsatile flow and was calibrated ∼1.45 cm upstream of the test valve site. The Projected Open Area (POA) was measured by a photodiode which was calibrated at the focused image of the telecentric lens.

### Valve leakage measurement

#### Leak rate and effective leak area

Fig. 3 illustrates elements in a system that measures the leak rate through test valves (Scotten and Siegel 2014). Our results infer shear rate damage to formed blood elements and resultant persistent thrombogenic response that constitute a fluid mechanical basis for observed thrombogenic rates unique to individual prosthetic valves now broadened to include observations of TAVI-related thromboembolic events. Our methodology enables resolvable regional flow velocity and focuses on overt flow transients previously overlooked and hidden due to brevity, small paravalvular spaces and instrumentation limits. We hypothesize that elevated flow velocity transients during the closing phase of the cardiac cycle may represent a primary factor in TAVI thrombogenesis particularly when trivial PVL is present and/or with greater aortic insufficiency (AI) within calcified and tortuous backflow pathways. Bioprosthetic valve leaflet closure, assisted by the flow deceleration phase, reduces transvalvular pressures and volumetric backflows, leading to the lower thrombogenic potential associated with these valves relative to MHVs. However, when PVL is present (e.g., after TAVI procedures), pathologic backflow velocity transients can be present.

**Figure 1.**
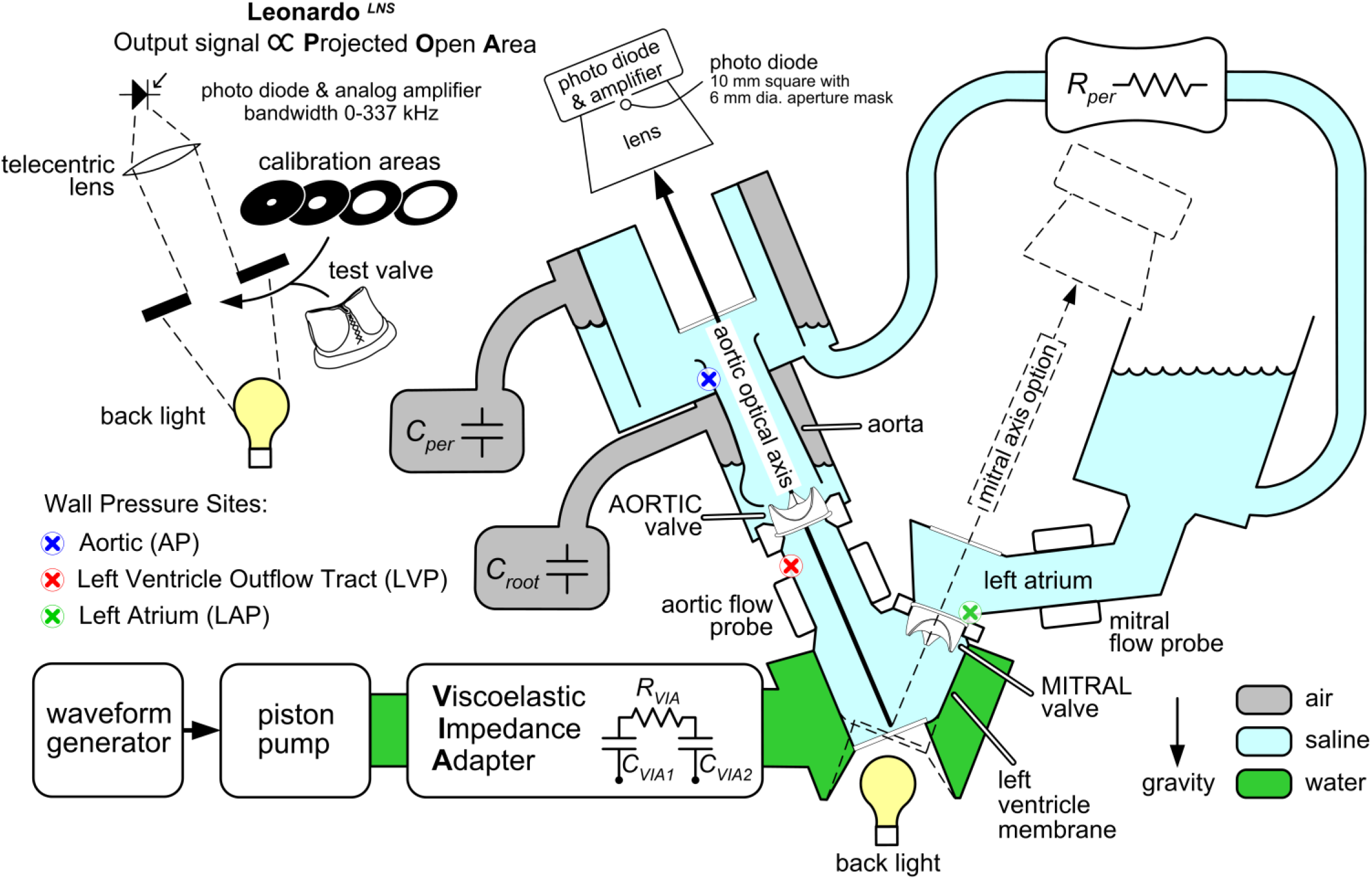
Leonardo^LNS^ pulse duplicator system opto-electronic measurement of valve Projected Open Area (POA) hydrodynamic simulation uses reduced models of Compliance and Resistance: C_VIA1_ 120 mL; C_VIA2_ 50 mL; C_per_ 615 mL; C_root_ 640 mL R_per_ adjusted for normal mean aortic pressure of 100 mm Hg; R_VIA_ 200 c.g.s. units. test fluid saline; cardiac output ∼ 5 L/min. VIA, Viscoelastic Impedance Adapter, model SPH3891B (ViVitro Labs Inc. Victoria, Canada Table 1 lists valves tested in our study including controls. The best closure performance is for valves iValve1.1 and iValve9.0 shown in outflow views in Fig. 2.

**Table 1.**
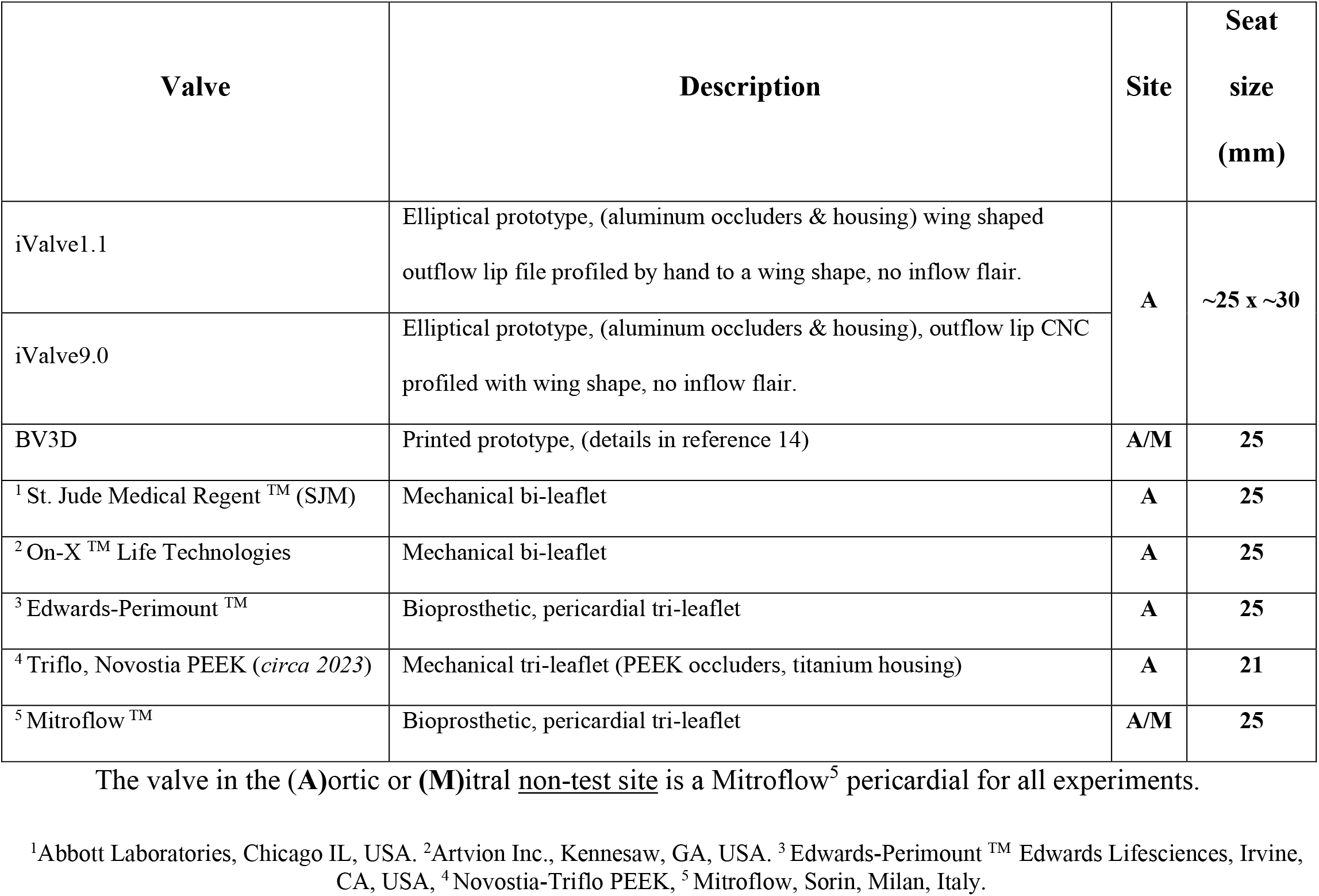
Test valves and controls.

**Figure 2.**
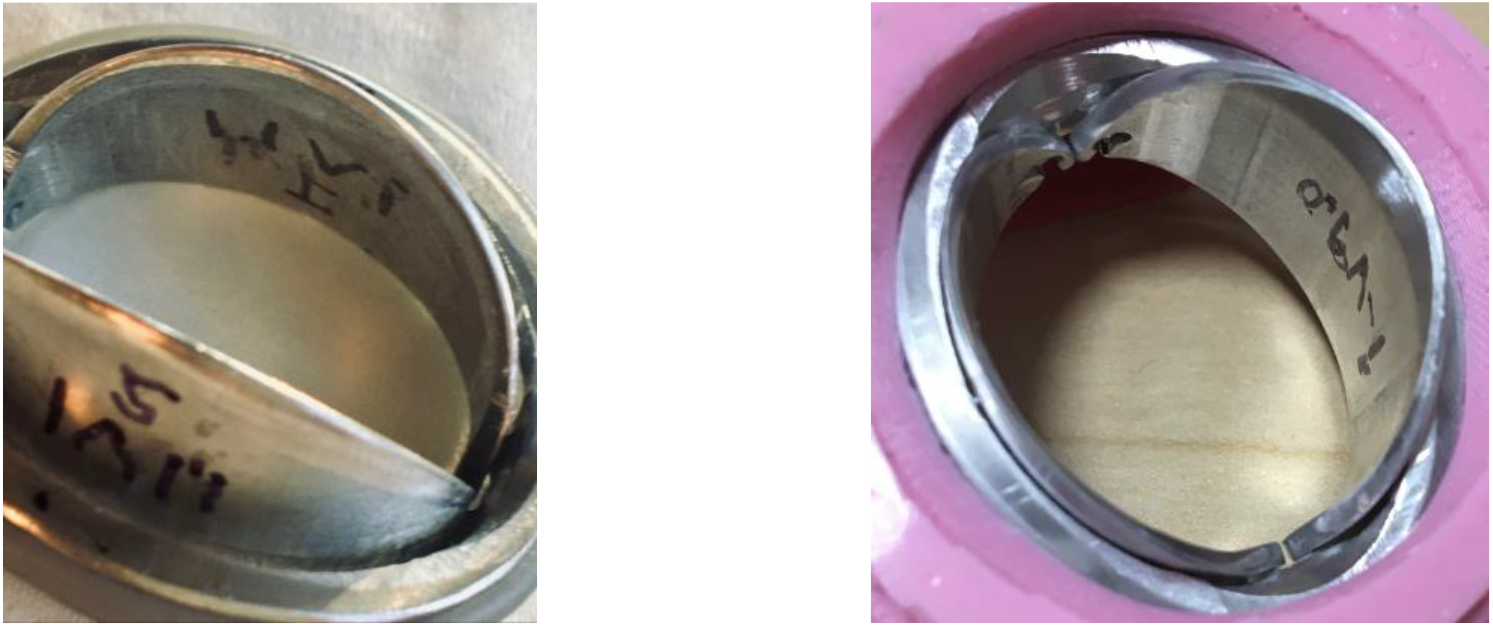
Outflow photo views of elliptical aluminum prototype mechanical valve models iValve1.1 (left) and iValve9.0 (right) that exhibit favorable closing dynamics.

**Figure 3.**
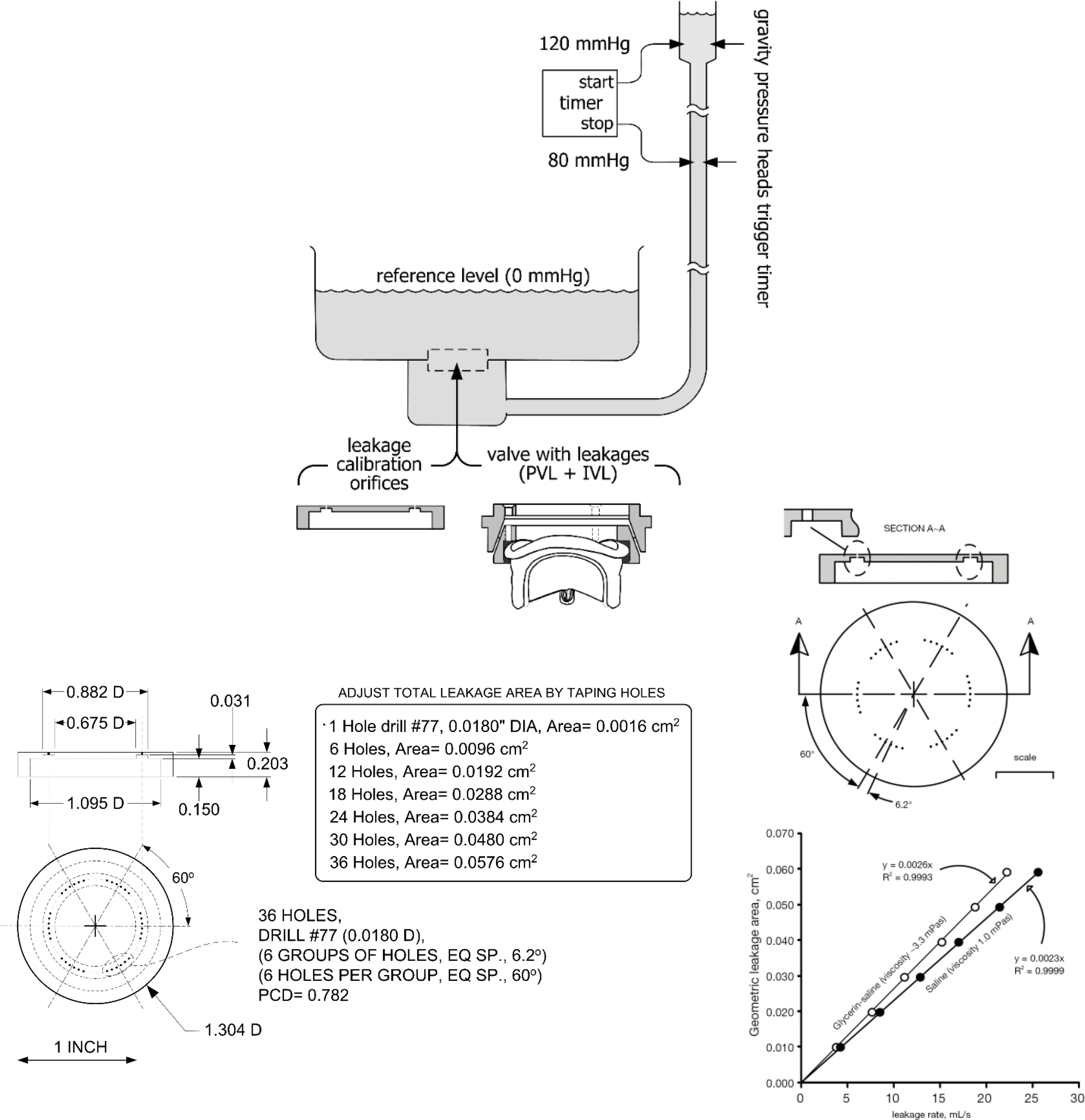
Valve leakage tester utilizes gravity pressure head and purpose-interfaced stopwatch timer. Lower panels show multiple orifice area specifics applicable to calibration data.

#### Statistical analysis

We report valve closure average velocity measurements and cycle-to-cycle variations of the 10 negative peak flow velocities to a 95% confidence level (CL). An EXCEL** data analysis tool labelled descriptive statistics is utilized together with the options -Analysis Tool Pak and -Solver Add-in.

## RESULTS

The SJM was the first all-carbon valve implanted in 1977 (Emery et al. 1979). The improved hemodynamics of the SJM influenced the On-X housing length, inflow profile and leaflet retention scheme intended to decrease anticoagulation requirements in low-risk patients (Wang et al. 2023). These valves have long published histories of wide clinical application for both aortic and mitral replacement and are standard-of-care devices.

Kinematic and hydrodynamic profiles for several valves are shown in Fig. 4. In Fig. 5. Negative peak flow velocities and CL = 95% indicated by CL bars adjacent to the flow velocity waveforms show the predicted uncertainty. A summary of valve peak flow velocities is shown in Fig. 6, which includes previously published data for a printed prototype-valve BV3D. All tested valve results may be compared to control valve data presented in Fig 7.

**Figure 4.**
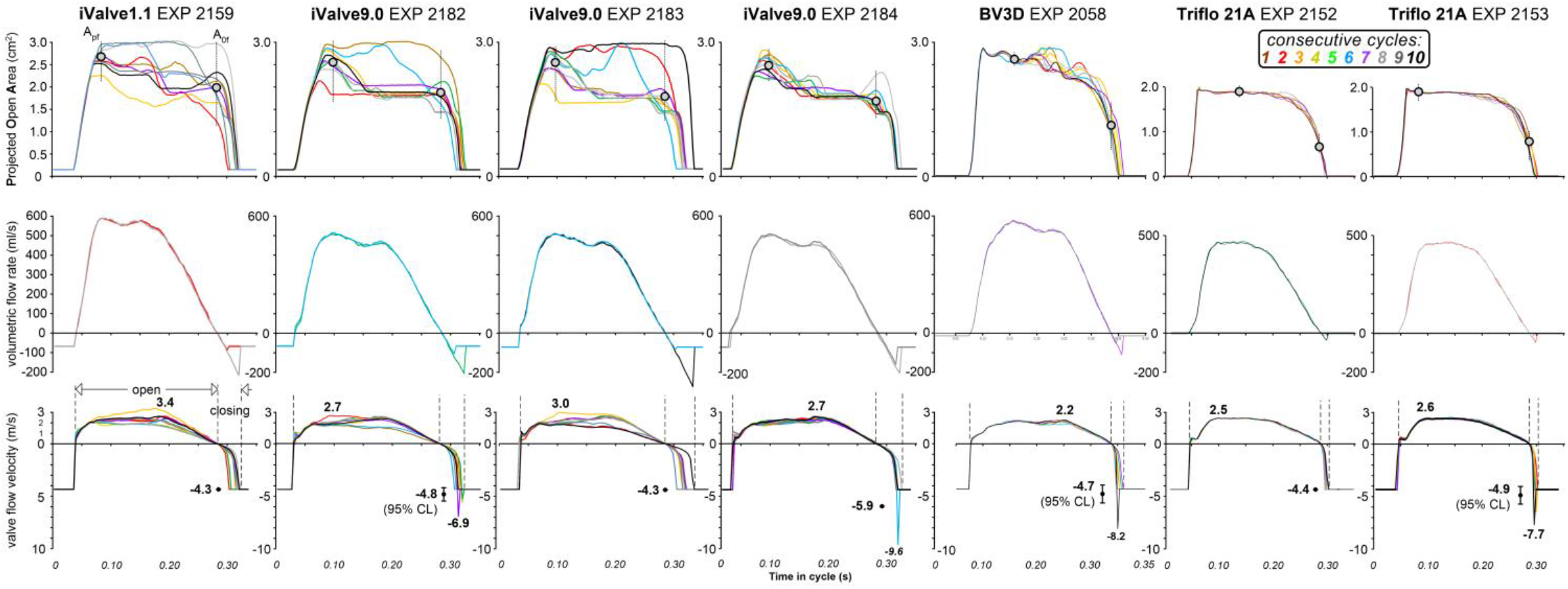
Mechanical aortic valves include aluminium proto-valve iValve9.0 designed and CNC machined by co-author (DG). The influence of small outflow edge details on iValve opening and closing performance was assessed. A_pf_ denotes average POA at peak aortic flow whereas A_0f_ denotes average POA at zero aortic flow. Experiment repeatability was assessed by waveform similarity in three experiments recorded over a ∼5-minute period (e.g., EXP 2182, 2183, 2184). The mean closure flow velocity range for valve iValve9.0 measured (4.3-5.9 m/s), indicative of superior closing performance with reduced thrombogenic potential relative the control valve flow velocity range (6.0-15.2 m/s). Valve closure response is benefited by iValve aerofoiled occluder outflow edges. In the lower panel, absence of “CL” whisker lengths is a notable signature of soft closure. **Microsoft, Redmond, WA, USA

**Figure 5.**
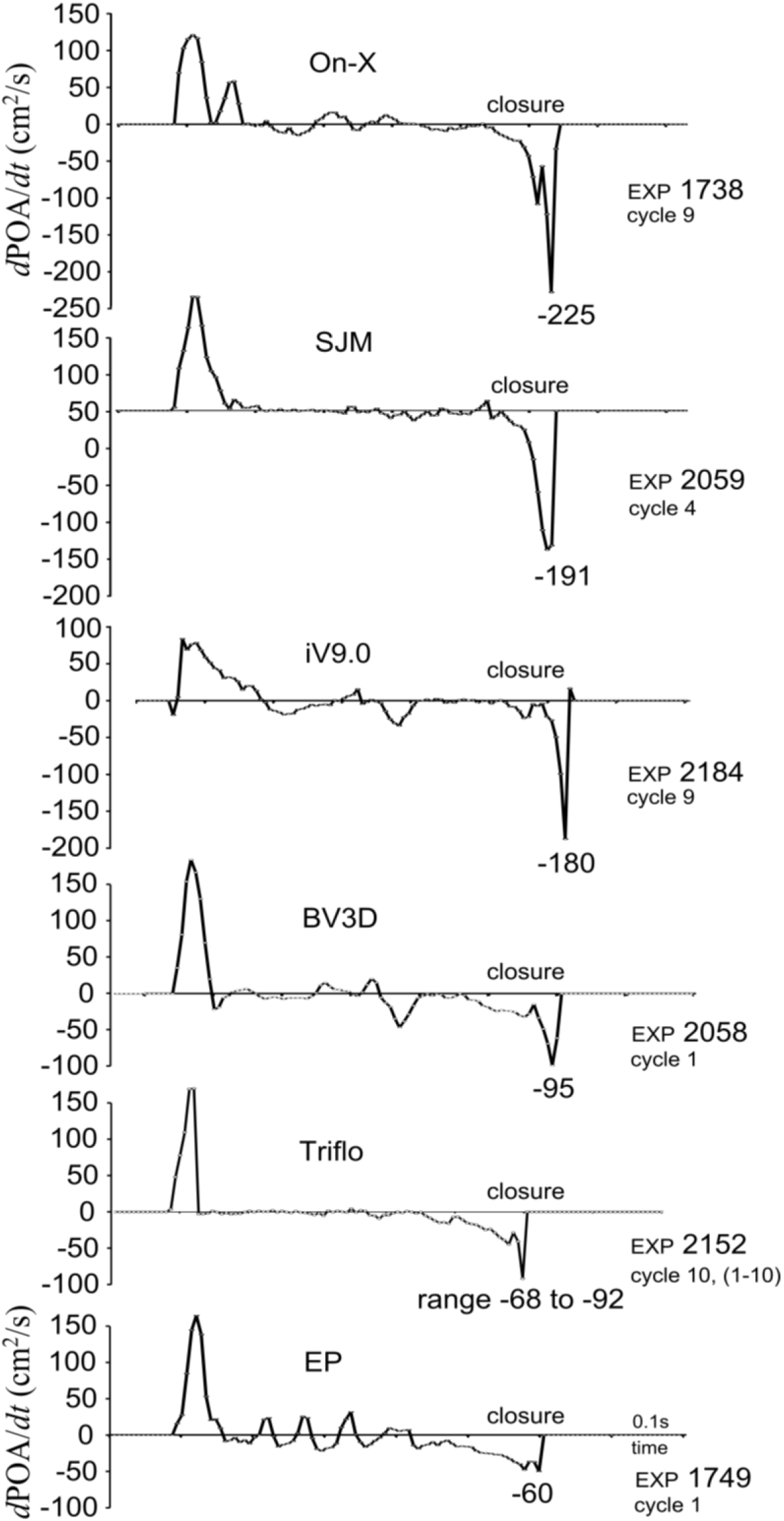
Shows **t**ime-dependent projected open valve area (*d*POA/*d*t). Low magnitude negative peaks signify moments of “soft closure”. The EP (and controls BV3D, iV9.0) exhibited soft closure in contrast to the SJM and On-X valves.

**Figure 6.**
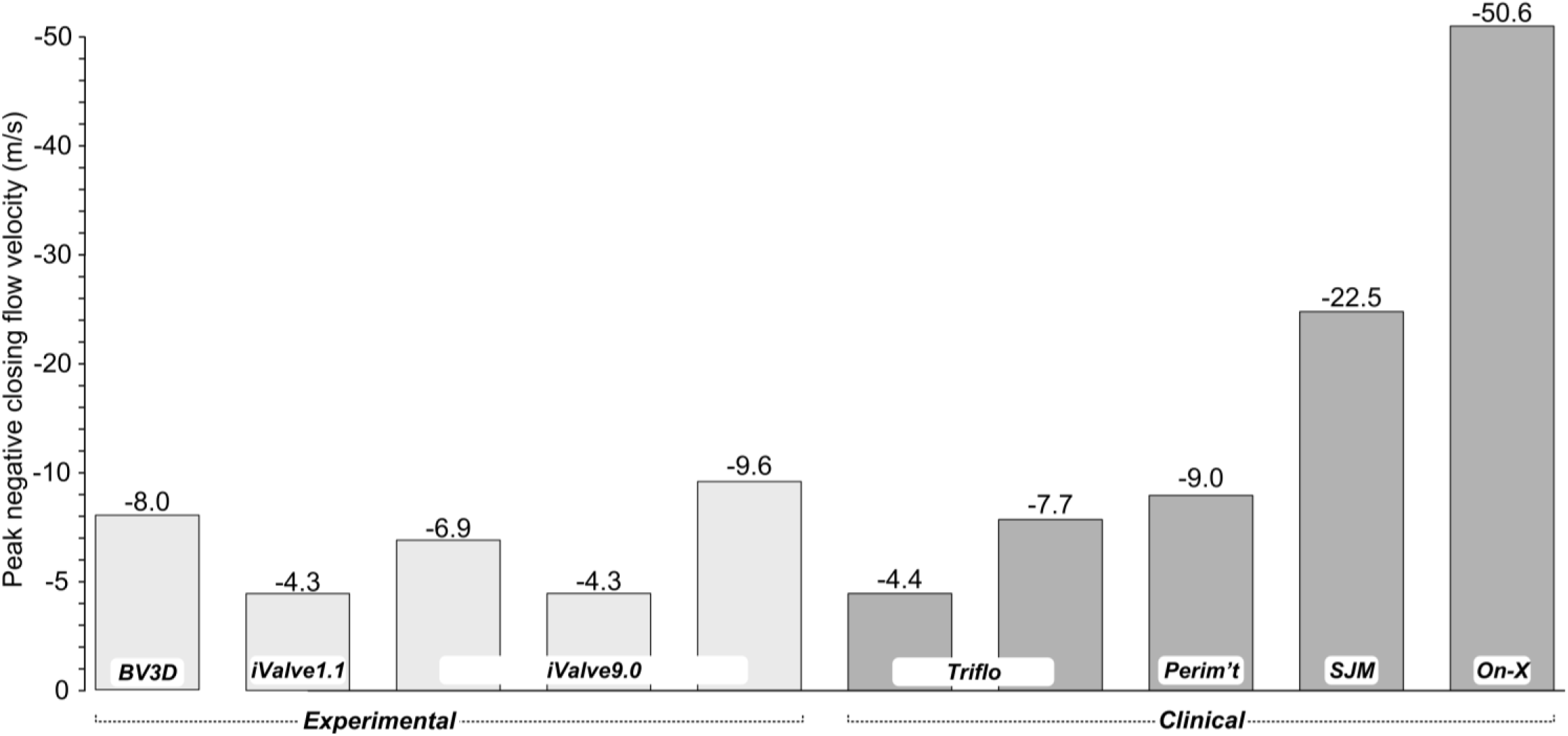
Summary bar graph of peak negative closing flow velocities for experimental and clinical (control) valves. BV3D is a printed prototype valve crafted by co-author (LNS) to study occluder geometry and pivot location effects on dynamic behavior. The Triflo features a valve seat size of approximately 21 mm, whereas the other valves have a seat size of 25 mm.

**Fig. 7.**
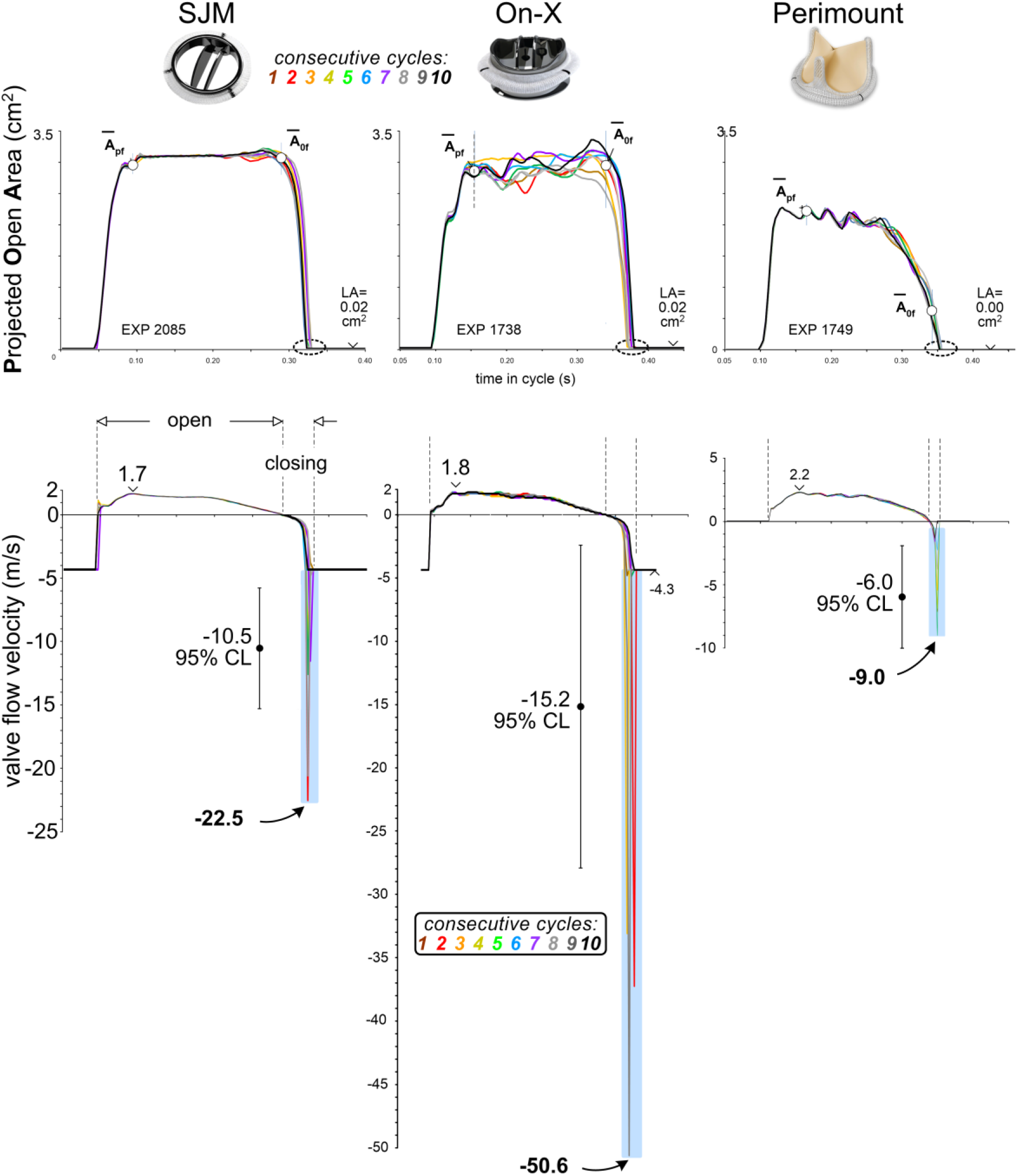
Control valves POA and flow velocity waveforms. Time-dependent flow velocity equals volumetric flow rate divided by valve Projected Open Area (POA). Dashed ovals on POA waveforms indicate valve closure moments. Regions highlighted in blue are where shock water hammer and transient negative flow velocities emerge.

The Perimount pericardial valve was chosen as a control, since it has demonstrated anticoagulation independence in numerous patients. Notably, the Perimount, iValve1.1 and iValve9.0 perform with minimal or absence of flow velocity spikes. Thus, the prototype valve designs with unique “soft closure” dynamics are evidence for reduced blood shear damage potential by exhibiting lower *d*POA/*d*t values relative to the MHV control valves. In vitro results seen in Fig. 5, support the conclusion that mechanical valves have closing response profiles with low *d*POA/*d*t values exemplify potential for anticoagulation independence. While the experimental bi-leaflet iValves and the tri-leaflet Triflo PEEK valve have dissimilar number of leaflets and leaflet retention (pivot) geometry, they share beneficial closure responses and flow velocity profiles with the pericardial control valve. As the Triflo PEEK valve is currently under clinical trial, early indications of low-no anticoagulation behavior may emerge shortly. Closing phase dynamics and the shear force damage to blood cells are implicated in the thrombogenic response to implanted valves and associated anticoagulation requirement.

## DISCUSSION

Motivated by the unmet need for a surgical mechanical valve implant with anticoagulant independence, we tested in vitro several innovative valve designs and compared to control valves and empirically fine-tune closure performance to minimize potential blood damage due to fluid shear forces. This was consistent with our intention to challenge the longstanding design bias that focused exclusively on forward flow detail and ignored valve closure aspects that we suspect play a key role in valve-related thrombogenicity.

## CONCLUSIONS

Slight differences in outflow and inflow edge configuration of prototype iValves have a profound impact on closure performance. This finding revealed that minor dimensional details are important, and valve geometry must be accurately fabricated and studied. Multiple design iterations may be required to empirically optimize valve geometry. In this regard, the study of the laboratory-modified valve (iValve1.1) provided great insight and appreciation for occluder design detail that has a measurable impact on closure dynamics, shear and potential thrombogenicity. This confirms that attention to minor detail is consequential in valve design.

The results presented here challenge the longstanding biases in valve design and highlight the importance of closure behavior, as evidenced by the integration of valve kinematic and hydrodynamic measurements. Testing a variety of unique experimental valves has opened a pathway for the development and introduction of advanced mechanical valves. Achieving anticoagulation-free mechanical valve performance, often considered the “Holy Grail,” may now be within reach.

## Data Availability

All data produced in the present study that is not presented in the manuscript are available upon reasonable request to the authors.

## Conflict of Interest Statement

Authors LNS, RS, DB, JWD report no conflicts of interest.

## Contributions

(I) Methodology, conception, and design: LNS; (II) Project administrative support: LNS; (III) Provision of study materials: LNS, DG; (IV) Collection and assembly of data: LNS; Data analysis, interpretation, validation: LNS, DJB; (VI) All authors have contributed.

## Acknowledgments

Trevor Scotten’s help with iValve experiments is appreciated.

## Competing Interests

None declared.

## Financial Disclosure (per coauthors HM, DG)

University of British Columbia, Mitacs Accelerate, Vancouver, Canada, Karl Im, Angeleno Medical LLC, LA, USA.

## Data availability statement

The core data supporting this study are available from the corresponding author, [LNS], upon reasonable request.

## Open access

Anyone can share, reuse, remix, or adapt this material, provided commercial purposes are not associated and the original authors credited and cited.

## CONTINUES WITH SUPPLEMENTAL MATERIAL…

**Figure S1.**
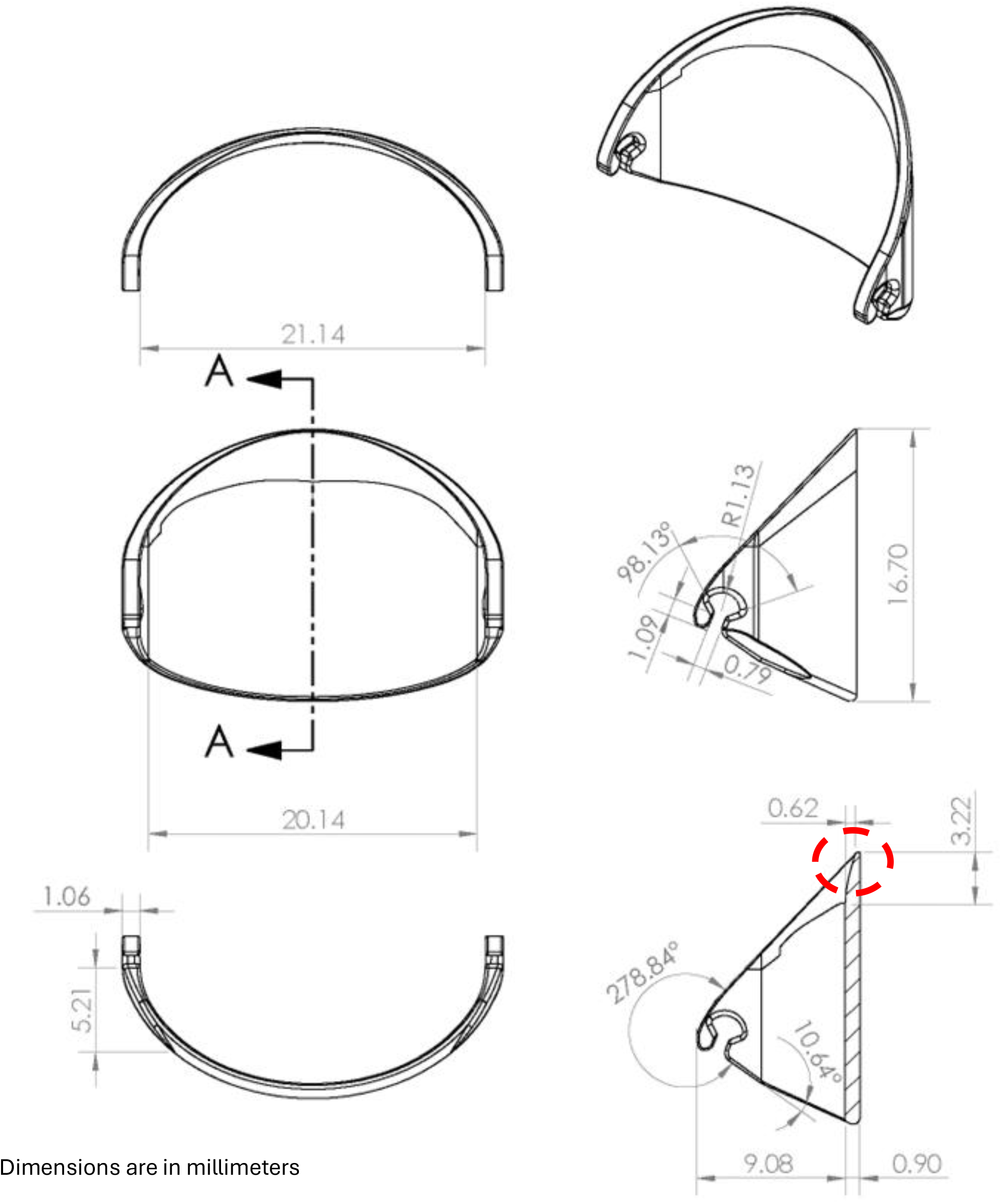
Sketches of prototype valve iValve9.0. Leaflet dimensions highlighted by the dashed red circle influence the beneficial soft closure response.

## Notes

### Competing Interest Statement

Financial disclosure per coauthors HM and DG: University of British Columbia, Mitacs Accelerate, Vancouver, Canada, Karl Im, Angelo Medical LLC, LA, USA.

### Funding Statement

This study was funded by University of British Columbia, Mitacs Accelerate, Vancouver, Canada, Karl Im, Angeleno Medical LLC, Los Angeles, USA

### Summary of Updates

Figure 4 ands 7 now includes color coded consecutive cycle legend insert. Minor text.pdf pagination format adjustments.

